# Abnormalities of the rib growth plate and the periphysis of previously healthy infants and toddlers dying suddenly and unexpectedly

**DOI:** 10.1101/2020.06.25.20139832

**Authors:** Elisa Groff, Suma Uday, Rita Zapata Vázquez, Khairul Zainun, Marta C Cohen

## Abstract

**Aims:** Histological examination of the rib is of critical value in perinatal pathology and points to the health of the child preceding death. The rib is considered ideal because it is the most rapidly growing long bone in infants and demonstrates growth arrest at onset of the insult. We aimed to identify: 1: changes in the perichondrial ring in the rib of infants and children up to 16 months of age dying suddenly at our institution and 2: any association with presence of histological changes of vitamin D deficiency (VDD) /metabolic bone disease (MBD) in the growth plate.

**Methods:** Retrospective review of the perichondrial rib histology and comparison with the presence or absence of histological features of VDD in the growth plate of 167 cases. The cases were anonymised and divided in six age/sex categories.

**Results:** Periphyseal abnormalities were only seen in 38% of the cases; of whom 33% had established and 67% had mild changes. Only 14.5% of cases with established histological appearance of VDD at the growth plate had significant PR abnormality; of whom majority (83%) were ≤3 months of age and none ≥9 months old, reflecting a temporal relation with birth and beyond the perinatal period.

**Conclusion:** The histological changes in the perichondrial ring are significantly associated with histological changes of VDD /MBD at the rib growth plate with an Odds Ratio of 3.04.

## INTRODUCTION

Histological examination of the rib is of critical value in perinatal pathology and points to the health of the child preceding death. The rib is considered ideal because it is the most rapidly growing long bone in infants and demonstrates growth arrest at onset of the insult (1-4). The post mortem (PM) examination protocol in our institution includes the removal of the right 5^th^ and/or 6^th^ rib as a means of assessing the child’s previous health; a practice which was introduced by the late John Emery (2,3). Most studies reporting changes at the costochondral junction (CCJ) have focused on the epiphyseal or metaphyseal changes and not on the periphysis. This is a fibrochondro-osseous structure that encircles the metaphysis and the adjacent CCJ of the tubular bones in infants and young children (5-7). It consists of a wedge-shaped cellular zone (groove of Ranvier) that surrounds the cartilaginous portion of the growth plate (or physis) and a thin layer of intramembranous bone (bone collar, bone bark or perichondrial ring of LaCroix) (6-8). See figure 1. Histologically, the Ranvier and LaCroix zones are a single structure depicting the thin layer of intramembranous bone at the periphery of the growth plate and metaphysis. The fibrous zones and Ranvier and LaCroix are richly supplied with blood from several perichondrial arteries, contrasting with the naturally avascular hypertrophic zone of the fully developed growth plate (6). The principal effect of the periphysis is on the metaphyseal collar (9): the groove of Ranvier is wedge-shaped collection of cells which provides chondrocytes for the longitudinal growth of the growth plate or physis and the perichondrial ring (PR) provides mechanical support to it (6, 8, 10). PM histological abnormalities at the rib growth plate in vitamin D deficiency (VDD) /metabolic bone disease (MBD) have been well defined by us (11) and other reports alike (12, 13). The changes reported to date have however focused on the epiphysis and metaphysis but not the periphysis as we mentioned earlier. Infants and young children with VDD show histological changes in the growth plate (and in the trabeculae) undergoing *endochondral ossification*. Concurrent features in the periphysis, which undergoes *intramembranous ossification*, have not been described. We hypothesize that young infants with histological features of VDD/MBD present in the growth plate (11-13) will associate with a simultaneous thickening of the periphysis (groove of Ranvier and PR of LaCroix). We envisage this phenomenon as a compensatory mechanism to stabilize the metaphysis.

**Figure 1:**
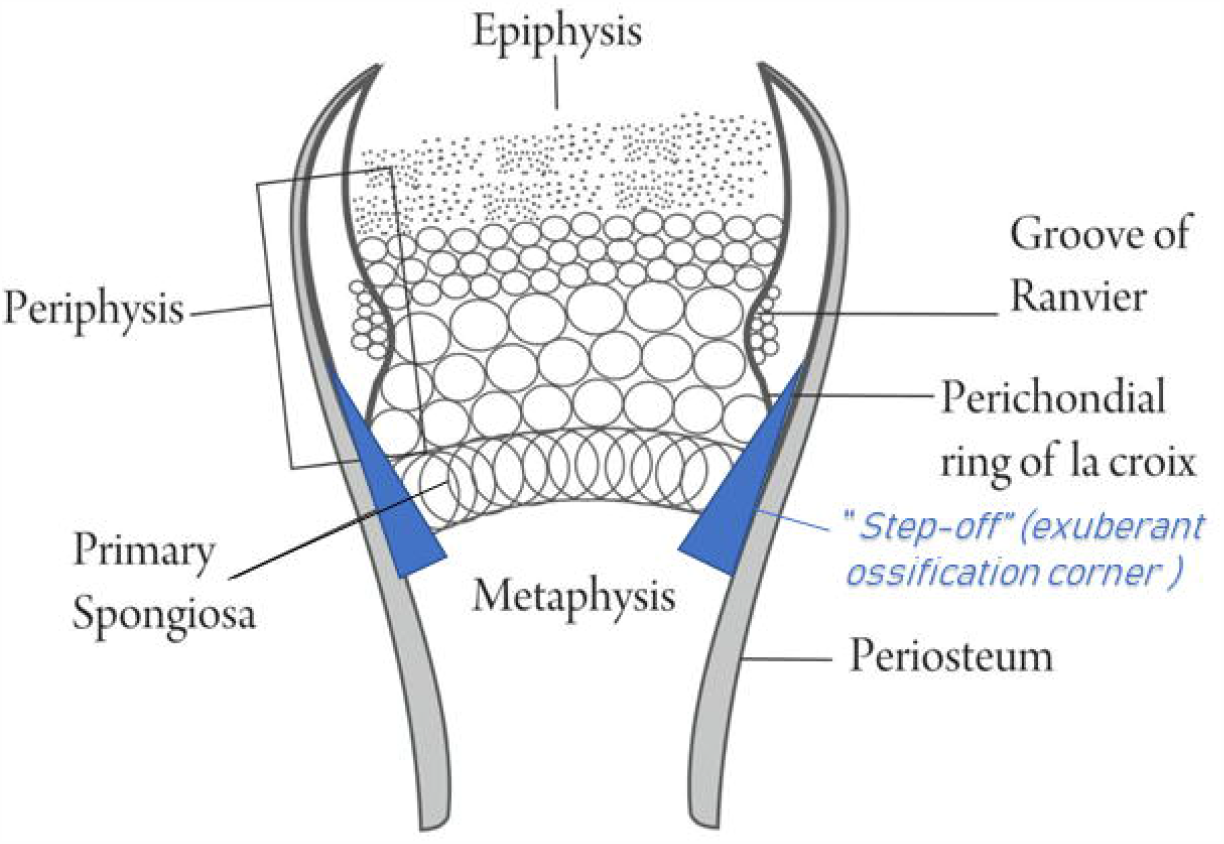
Schematic drawing of the periphysis, encircling the metaphysis and depicting the wedge-shaped groove of Ranvier and the thin layer of intramembranous bone (bone collar, bone bark or perichondrial ring of La Croix)

The aims of our study are to identify: 1: changes in the PR at the CCJ of the rib of infants and children up to 16 months of age dying suddenly and unexpectedly at our institution and 2: a possible association between the histological changes in the PR with the presence of histological changes of VDD/MBD at the growth plate (11).

## MATERIAL AND METHODS

### Design

Retrospective study

### Subjects

Infants and children ≤16 months of age undergoing PM examination, during a 9-year period, at a single regional centre for sudden unexpected death.

### Data gathering

Anonymised data for all patients with consent was collected from the institutional PM data base. The search terms included: sudden infant death syndrome (SIDS), undetermined, unexplained, unascertained, or sudden unexpected death in childhood. Data collected included age at death, gender, serum 25 hydroxy vitamin D (25OHD) concentrations (where available) and cause of death at PM.

The cases were anonymised and divided in six age/sex categories as follows: perinatal (between birth to 28 days), 1-3 months, 3-6 months, 6-9 months, 9-12 months, 12-16 months. Whenever an age-at-death-value was between the end- and the start-value of two categories, it was placed in the subsequent crescendo category, e.g. a baby who died at age of 3 months belongs to the age category 3-6.

### Description of abnormal rib histology

In each case the histology of the rib growth plate was reviewed and agreed by two paediatric pathologists (KZ and MCC). The presence or absence of histological features that characterise VDD was recorded (11-14): chondrocyte hyperplasia at the growth plate, woven bone in the trabeculae, penetrating vessel into the growth plate and widening of the CCJ with bulbous appearances on low magnification. Figure 2 a-d. The histological appearance of VDD was considered “established” when ≥ 3 features were present or “mild” when <3 features were identified.

**Figure 2:**
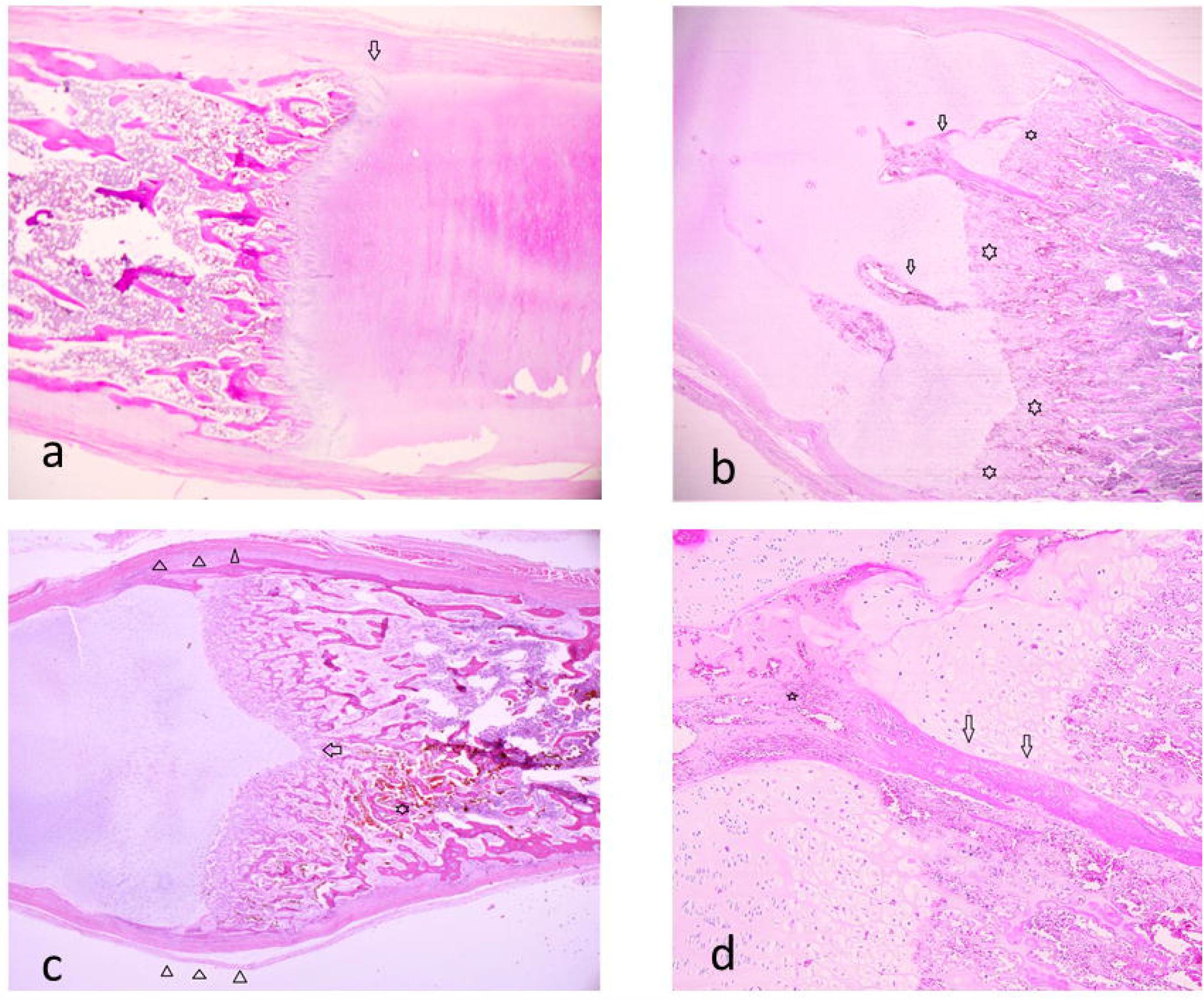
a: low power of a normal growth plate in a SIDS case; note the orderly columns of chondrocytes, without a bulbous shape or penetrating vessels and thin perichondrial ring (arrow) (H&E x 20). b; low magnification of the growth plate in a 2 months-old SIDS case with features of vitamin D deficiency as suggested by the irregular growth plate (arrow), with hyperplasia of chondrocytes, bulbous shape (triangles) and a fresh microfracture (star) (HE x 10); c: the growth plate in a 3 weeks old previously healthy SIDS case showing an irregular growth plate (stars), with bulbous shape and penetrating vessels (arrows) (H&E x 20); d: higher magnification of “c” showing ossification (arrows) of the abnormally penetrating vessels (star) into the growth plate. Note the irregularity of the columns of chondrocytes (HE x 40).

### Description of abnormal PR histology

The following features were recorded in order to assess the appearances of the PR: widening of the bark bone, exuberant ossification at the periphyseal corner (“step-off”) (5, 15,16) and/or abnormal metaphyseal collar. Figure 3a-d. The abnormalities of the PR were graded as: normal, mild, established.

**Figure 3:**
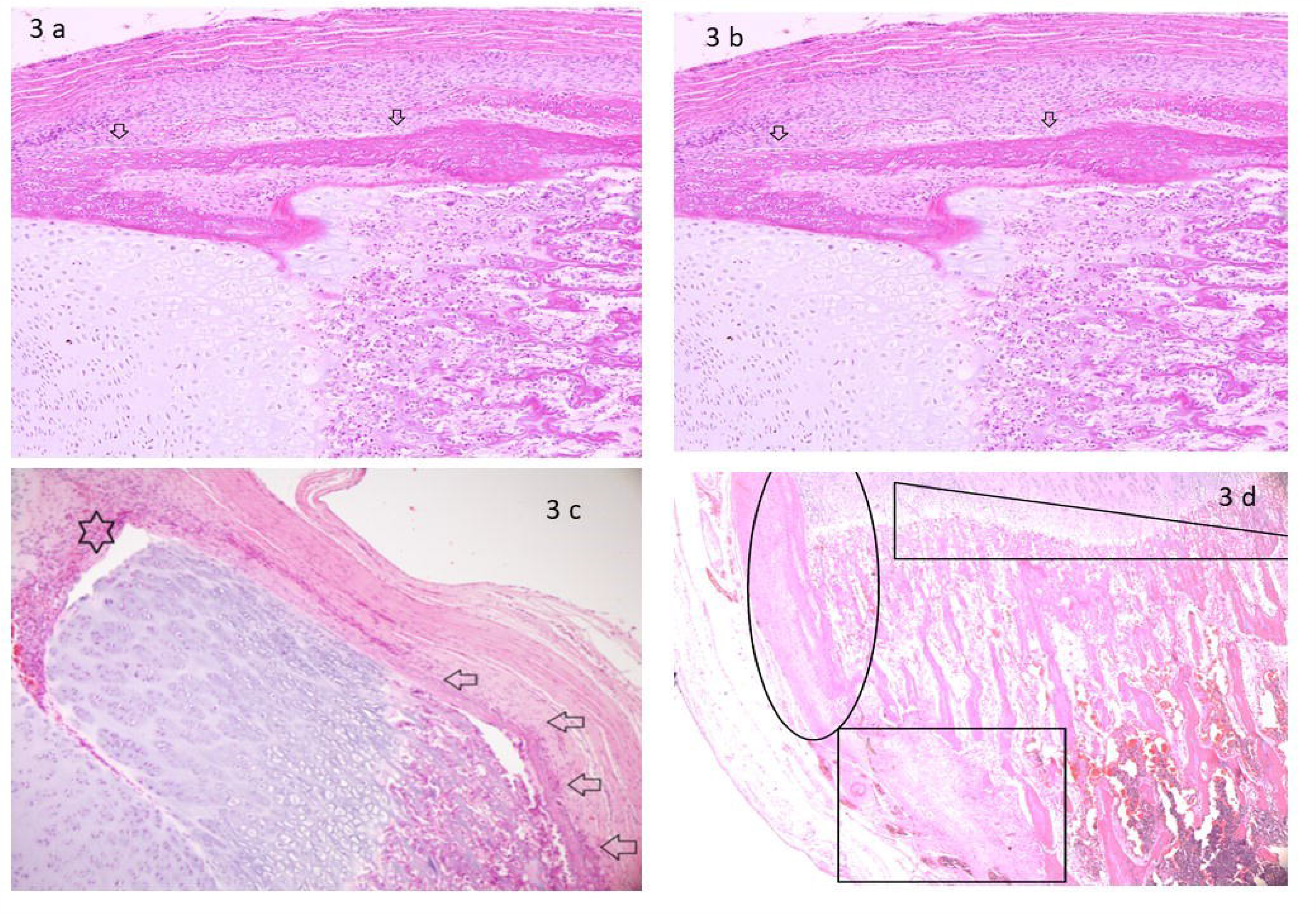
a and b: prominent perichondrial ring (arrows) in two SIDS case with features of vitamin D deficiency in the growth plate (corresponding to same patients shown as 2b and 2c respectively. H&E x 40); c: SIDS case with features of vitamin D deficiency in the growth plate and prominent perichondrial ring (arrow). Note a blood vessel penetrating the growth plate from the perichondrial ring (star) (H&E x 40); d: irregular growth plate with hyperplasia of chondrocytes (triangle area), prominent perichondrial ring (rectangle area) and abnormal periosteal reaction (oval area) (H&E x 20).

### Serum 25 hydroxy vitamin D measurement

Serum 25OHD was measured at a regional laboratory (University Hospitals Bristol, Bristol, United Kingdom) using ultra performance Liquid Chromatography tandem mass spectrometry.

For the purposes of this analysis the category stated in the report received from the laboratory was used: a 25OHD level of < 25 nmol/L (severely deficient); 25 −50 nmol/L (deficient), 50– 75 nmol/L (suboptimal); and >75 nmol/L (adequate).

### Statistics

We illustrate the changes in the PR at the CCJ of the rib using descriptive statistics, by age groups and by sext. The association between PR changes and histological features of VDD at the growth plate was evaluated with the chi-square test (𝒳^2^). The association between PR changes and serum 25OHD concentrations was assessed with the Fisher-test, considering that the sample was small and, thus, one of the assumptions of 𝒳^2^ test was not met (expected value in any box should be greater than 5). Further, we calculated the corresponding Odds Ratio (OR) as a quantitative measure of the associations. All tests were performed using the Statistical Package S-Plus ® v6.21, and Microsoft Excel-Microsoft Office 365 for the graphics.

### Consent and ethics

As per the Human Tissue Act, only those cases where there was consent to participation in research were included (19). The study was approved by the Local Ethic Committee (SCH/13/027).

### Funding

The study did not have any funding source.

## RESULTS

A total of 167 (101 males, 66 females) cases of sudden unexpected death were identified. The sex and number of cases per age group is shown in Figure 4.

**Figure 4:**
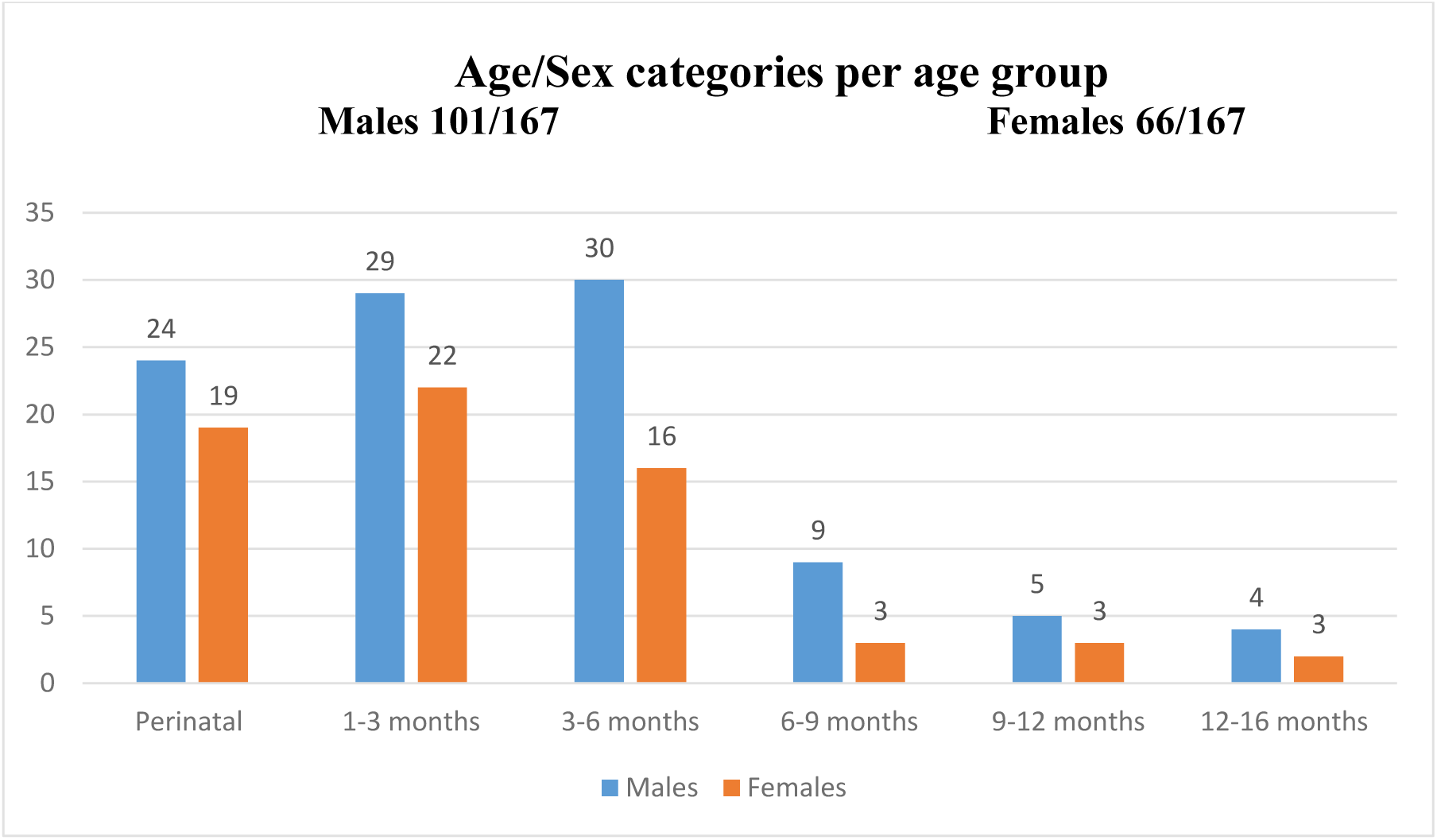
Cases distributed according to sex and age Cohort Age/Sex categories.

Histological appearance of the rib was normal in 21% (35/167), of which 22.8% were males (23/101) and 18.8% females (12/66). The rib growth plate was abnormal in 79% (132/167) of the cases, of which 58% were male (77/132) and 42% female (55/132). Of these, histological features of VDD/MDB were clearly established in 63% (83/132) of cases, 60% males (50/83) and 40% females (33/83), and were mild in 37% (49/132) of cases, 55% of which were male (27/49) and 45% female (22/49). Despite the higher number of males than females with sudden and unexpected death, the incidence of histological features of VDD was similar in both sexes. See Figure 5.

**Figure 5:**
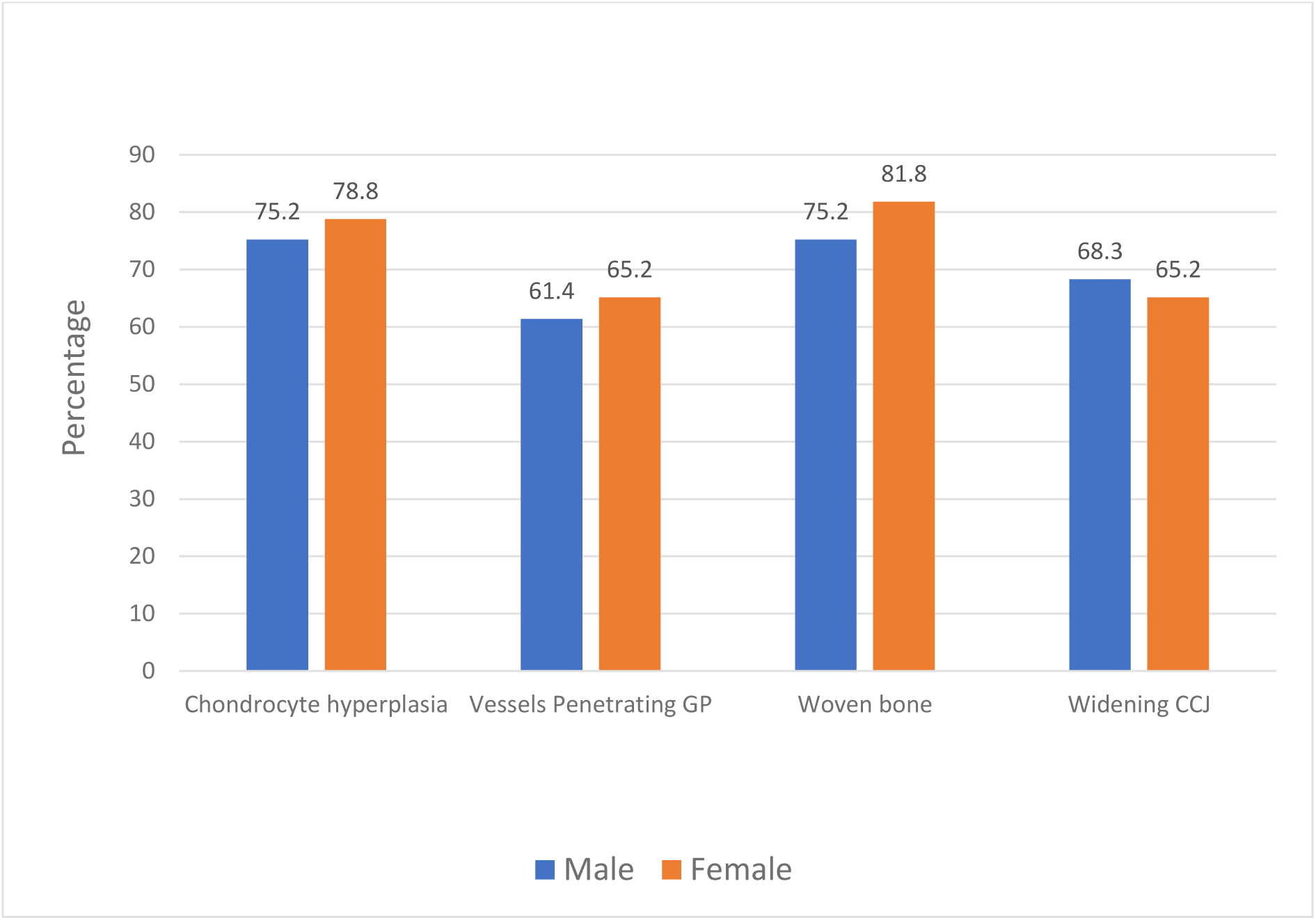
Distribution of histological features of vitamin D deficiency in the growth plate in each sex.

The PR was normal in 62% (103/167) of cases, 55% of which were male (56/103) and 45% female (46/103). The PR was abnormal in 38% (64/167) of cases, 73.5% of which were male (47/64) and 26.5% female (17/64). Of these, PR abnormalities were graded as mild in two thirds of cases (67%; 43/64, 28 males and 15 females) and established in one third of cases (33%; 21/62, 17 males and 4 females). The PR abnormalities in the 21 cases at the most severe end of the spectrum included: widening of the intramembranous layer of bone (bone bark), exuberant ossification at the periphyseal corner (“step-off”) and irregular metaphyseal collar. Figure 6a-b.

**Figure 6:**
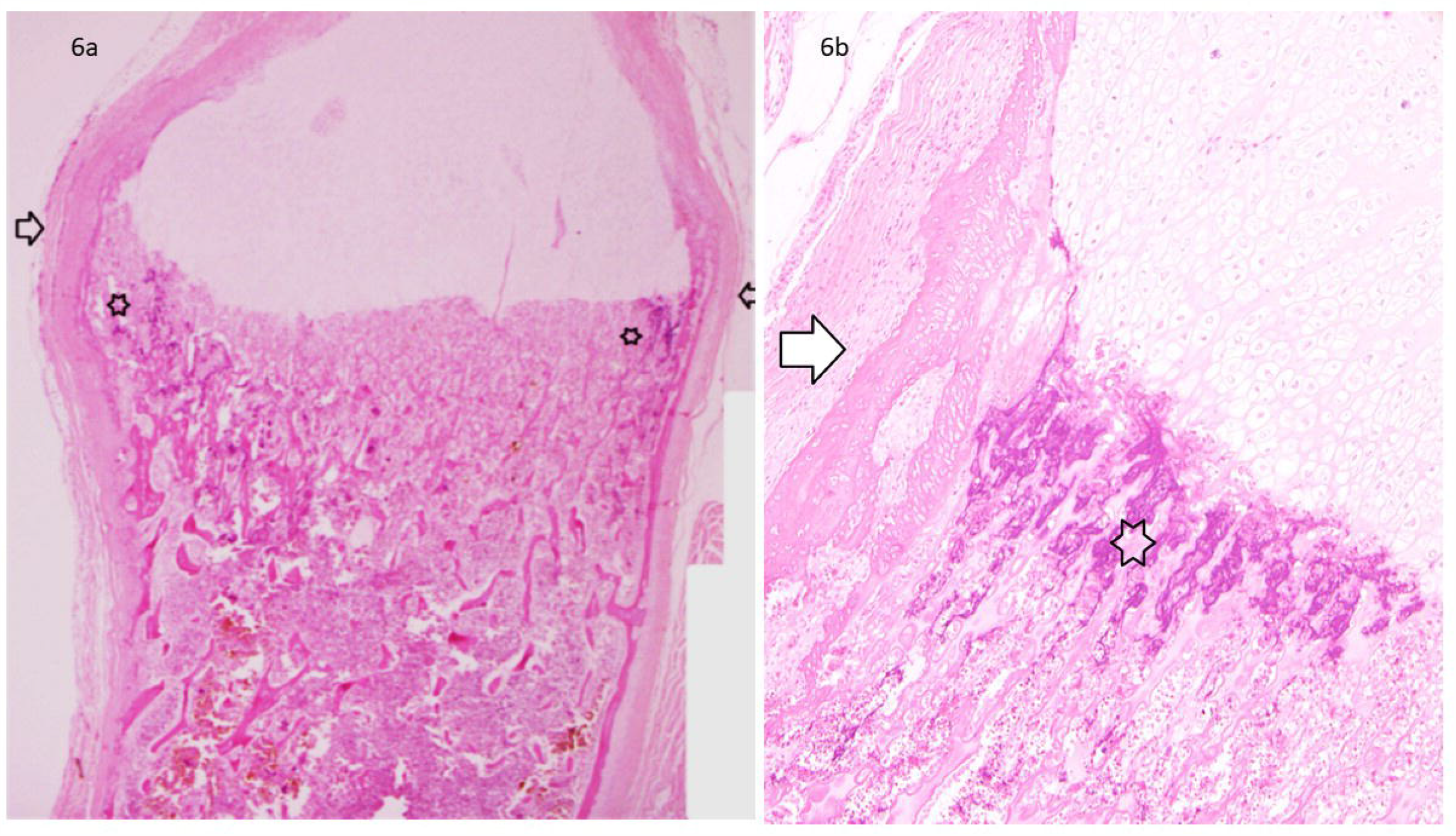
a: Low magnification showing a bulbous shape of the growth plate (arrows) with a prominent perichondrial ring and step off (exuberant ossification corner, stars) (H&E x 20); b: the abnormally prominent step off also shows exuberant mineralisation (star), which may should not be interpreted as a corner metaphyseal lesion (CML).

The presence of histological features of VDD at the growth plate show a significant association with the presence of histological abnormalities at the PR (𝒳^2^ = 5.35, df=1, *p* = 0.021); with an OR of 3.04 (95% CI: 1.24-7.45). These results demonstrate that the presence of histological features of VDD/MBD at the growth plate carries a 3.04 risk of PR abnormalities and vice versa (Table 1). Overall, of the 35 cases with a normal histological appearance of the growth plate, the PR was normal in 28 cases (80%) and abnormal in 7 cases (20%). Of the 132 cases with histological features of VDD/MBD in the growth plate, the PR was normal in 75 cases (57%) and abnormal in 57 (43%).

**Table 1:** Histological appearance of vitamin D deficiency/metabolic bone disease (VDD/MBD) and perichondrial ring features

|  | **Perichondrial Ring** |  |  |
| --- | --- | --- | --- |
| **Histological features of VDD/MBD** | **Abnormalities Established or Mild** | **Normal** | **Total** |
| **Established or Mild** | 57 | 75 | **132** |
| **Normal** | 7 | 28 | **35** |
| **Total** | **64** | **103** | **167** |

Serum 25OHD were measured at post mortem in 27/167 (21 M/ 6 F) cases. In this group, serum levels were adequate/suboptimal in 9 cases and deficient in 18 cases. The appearances of the PR in the 27 cases in which serum 25OHD was measured is shown in Table 2.

**Table 2:** Relation between vitamin D (VD) blood levels and the appearance of the perichondrial ring (PR).

| **VD level** | **Established or Mild abnormality of PR** | **Normal PR** | **Total** |
| --- | --- | --- | --- |
| **Severe deficiency or Deficient** | 13 | 5 | **18** |
| **Suboptimal or Adequate** | 2 | 7 | **9** |
| **Total** | **15** | **12** | **27** |

A close analysis shows that an abnormal PR was identified in 13/18 (72%) cases with serum 25OHD deficiency and in 2/9 (22%) cases with adequate/suboptimal serum 25OHD levels (see Table 3). In addition, in 15 cases with mild or established abnormalities of the PR, the serum 25OHD concentrations were deficient in 13/15 (87%) and adequate/suboptimal in 2/15 (13%) cases. There was a significant association between serum 25OHD concentrations and PR appearances (*p* = 0.037, Fisher test). Moreover, the corresponding OR value of 9.1 (95% CI: 1.4 a 59.6) suggests that the risk of PR abnormalities is nine times higher in the presence of VDD, and vice versa.

**Table 3:** Relation between vitamin D (VD) blood levels and the appearance of the perichondrial ring.

| VD level | HPR Normal | HPR Mild | HPR Established |
| --- | --- | --- | --- |
| Adequate | 5 | 0 | 1 |
| Suboptimal | 2 | 0 | 1 |
| Deficient | 3 | 2 | 7 |
| Severe deficiency | 2 | 1 | 3 |

HPR: histology of perichondrial ring; VD level: Adequate: >75 nmol/L; suboptimal: 50 – 75 nmol/L; deficient: 25 – 50 nmol/L; severe deficiency: <25 nmol/L; 27 cases = 21 Males/ 6 Females

## DISCUSSION

Our results demonstrate that nearly 80% of the cases presented variable histological changes of VDD in the rib growth plate; of whom 63% had established and 37% had mild changes. In contrast, periphyseal abnormalities were only seen in 38% of the cases; of whom 33% had established and 67% had mild changes. Only 14.5% of cases with established histological appearance of VDD at the growth plate had significant PR abnormality; of whom majority (10/12, 83%) were ≤3 months of age and none ≥9 months old. PR abnormalities are therefore less common than those of epiphysis and tend to be present at a younger age. Our results demonstrated that the presence of histological features of VDD at the rib growth plate show a significant association with the presence of histological abnormalities of the PR (𝒳^2^ = 5.35, df=1, *p* = 0.021); with an OR of 3.04 (95% CI: 1.24-7.45). Moreover, the presence of histological features of VDD at the growth plate had a 3.04 risk of histological PR abnormalities and vice versa.

Eighty four percent of the whole cohort (140/167; 83 males and 57 females) were ≤ 6 months of age, reflecting the current statistics for most victims of sudden and unexpected infant deaths in the United Kingdom which occur in the first 6 months of age, with a higher prevalence in males (20-22). Almost 50% (83/167) of our cases showed established histological evidence of VDD in the growth plate. The great majority of the cases (78%) were ≤3 months of age. Of the 18 cases with serological VDD, the majority (13,72%) also had histological evidence (established in 10 and mild in 3) while 5 (28%) cases had a normal histology. As in previous studies, this confirms a correlation between the histological appearance of VDD in the growth plate of the rib and the concentration of VD in serum (11,13).

Part of the role of the PR is to lay down a thin layer of bone through intramembranous ossification (bone bark), which provides mechanical support to the growth plate and the metaphysis, and increases the width of the physis (16, 18, 23,24). The intramembranous bone bark extends from the level of the proliferating chondrocytes in the epiphysis to the zone of primary ossification in the metaphysis (7). Our results showed that the PR abnormalities at the most severe end of the spectrum included: widening of the intramembranous layer of bone (bone bark), exuberant ossification at the periphyseal corner and prominent irregularities of the metaphyseal collar. An established abnormality of the PR was present in 12 of 83 cases (14%) with significant histological features of VDD at the growth plate (all ≤ 9 months with a peak between 0-3 months of age) and 4 of 35 cases with no histological evidence of VDD (11%) (all ≥3 months of age). The finding of abnormalities of the PR in cases with no evidence of VDD suggests that other metabolic pathways also likely influence the normal structure of the PR.

Notably, most infants with changes in the PR were <3 months of age which indicates that pre- and perinatal factors may also play a role. Studies of the CCJ in stillborn foetuses and newborns conducted in the ‘60s described a pattern of abnormalities similar to our findings (2,3). More recently also, placental abnormalities have been linked to abnormalities in the CCJ (4). Whilst vitamin D appears to be one factor, other factors that play a role in abnormalities of the PR are yet to be elucidated. Development of the foetal skeleton requires minerals such as calcium and phosphorus, acquired from the mother, and if the increased foetal demand in minerals is not met, then inadequate foetal bone mineralization may not occur (25-29). Vitamin D is transferred via the placenta predominantly as 25-hydroxyvitamin D, subsequently converted to 1,25-dihydroxyvitamin D in the foetal kidney (25). Health and disease in later life are partly determined in utero (30,31), and it has been proposed that it can affect structural, metabolic, physiologic, and behavioural development and modify response patterns that influence future susceptibility to diseases (32,33). This is relevant to bone structure, as maternal VDD at 18 weeks’ gestation has shown to be associated with lower peak bone mass among their children at 20 years of age (33,34). As expected, VD deposits in the newborn are related to maternal nutritional adequacy (34, 35). This correlation is maintained even 4 months after birth (36).

Experimental studies demonstrated that the cells from the groove of Ranvier are pushed into the reserve and proliferative regions of the growth plate in order to supply cells for the reserve layer of chondrocytes. This causes an expansion of the diameter of the growth plate and serves as cartilaginous stem cells, thereby playing a major role in endochondral ossification (23, 37). The Indian hedgehog (Ihh), member of the hedgehog family, is a key molecule coordinating these processes (38,39). Ihh is also part of the vitamin D pathway: it stimulates chondrocyte proliferation in the growth plate, it prevents chondrocyte hypertrophy and regulates bone formation in the perichondrial collar and in the trabecular bone below the growth plate (38, 39). These physiological processes will naturally be influenced by the vitamin D status.

The features identified in our study of the periphysis of infants with histological appearances of VDD have not been previously addressed. Much attention has been given to describe how differentiation of the hypertrophic chondrocytes in the growth plate and the subsequent calcification of the matrix are impaired in VDD leading to the flaring of the ends of the long bones (40). The bone bark in foetuses can be identified after 10 weeks’ gestational age, and at 16 weeks all the structures in the periphysis became clearly defined (7). It should be noted that the bone bark of the PR is more prominent in the developing bones in which there is extensive metaphyseal remodelling (37). At the metaphyseal end, the intramembranous bone bark is overlaid by the periosteum and at the same time in intimate contact with the cartilage matrix undergoing endochondral ossification in the zone of primary bone trabeculae (7,37).

The membranous bone produced by the periphysis shows variation in thickness in different parts of the long bones (7) and can adopt the appearance of a spur (or “step off”). It should not be mistaken for a child abuse fracture or be considered in itself a manifestation of rickets (5). The rapid bone growth and remodelling seems to be responsible for subperiosteal new bone formation, a common finding in infants between 1 and 4 months of age (41). During the active stage of rickets, the impaired mineralization of the zone of provisional calcification and the PR results in hypertrophy of the chondrocytes and accumulation of osteoid and un-mineralised matrix, which can be exuberant in some infants. Experimental studies in dogs have demonstrated VDD induced proliferation of exuberant cartilaginous tissue (42), and osteoid deposition in the exuberant cartilage and proliferation of capillary vessels in the healing stage of VDD can closely mimic a healing fracture to those unaware of this process. Similarly, the bone bark may be radiologically visible at the edge of the physis at the wrist of infants and children up to 12 ½ years and should not be mistaken for fracture or rickets (43).

The main limitation of our study is that serum 25OHD concentrations were measured in only a limited number of cases, nonetheless the focus of our study was to describe the periphyseal changes in relation to well described histological epiphyseal changes of VDD and not serum levels.

In conclusion: Epiphyseal changes of VDD/MBD at the growth plate are well established, and in this report, we describe the associated periphyseal changes which have not been previously described. Although less prevalent, the histological changes in the perichondrial ring are significantly associated with histological changes of VDD/MBD at the rib growth plate with an OR of 3.04. And such changes are widely prevalent in those under 3 months of age, hence future prospective studies should aim to explore not only the foetal but also maternal contributing factors.

## Data Availability

Data is available upon reasonable request

## Acknowledgment

The authors are grateful to Praveen A Lakshmipathi for his help with the drawing of figure 1.

